# An Ultrasensitive and Novel Gene Sensor to Monitor Gut Microbiome

**DOI:** 10.64898/2026.04.30.26352170

**Authors:** Muhit Rana, Meredith Stewart, Marinelle Rodrigues, Erdal Toprak, Andrew Koh, Avni A. Argun

**Affiliations:** Giner, Inc, 89 Rumford Avenue, Newton, MA 02466, USA; Department of Pharmacology, University of Texas Southwestern Medical Center, Dallas, TX 75390, USA; Lyda Hill Department of Bioinformatics, The University of Texas Southwestern Medical Center, Dallas, TX 75390, USA; Division of Hematology/Oncology, Department of Pediatrics, University of Texas Southwestern Medical Center, Dallas, TX 75390, USA; Harold C. Simmons Comprehensive Cancer Center, University of Texas Southwestern Medical Center, Dallas, TX 75390, USA; Department of Microbiology, University of Texas Southwestern Medical Center, Dallas, TX 75390, USA

**Keywords:** Multi-drug-resistant organisms (MDROs), antibiotics, gene, electrochemical, DNA, detection

## Abstract

Infections caused by multi-drug-resistant organisms (MDROs) pose a significant public health threat, responsible for over 2 million hospitalizations and 23,000 deaths annually in the United States. Microbiome dysbiosis (imbalance) is considered one of the main causes for MDRO colonization and the resulting infections. Rapid detection and intervention of MDRO outbreaks are crucial to alleviating strain on patients and healthcare facilities. Current diagnostic methods for MDRO detection are too slow and costly to provide the rapid MDRO detection necessary for patient care facilities. Here we present a rapid, accurate and cost-effective electrochemical sensor capable of MDRO detection down to ∼10^4^ colony forming units (CFU)/g in mice and human stool samples. Our novel sensor utilizes probe-modified <u>S</u>creen-<u>P</u>rinted <u>E</u>lectrodes (SPEs) capable of hybridizing target gene sequences associated with MDROs. The resulting probe/target complex generates a unique and highly sensitive signal detectable down to 10 atto molar or 10 CFU/mL of target TEM-1 gene. The use of these pre-functionalized SPEs reduces individual sample analysis time to less than an hour. Several target sequences from two chromosomal target genes (AmpC and AcrB found in *E. coli*) have been identified and successfully detected in clinical stool samples with results comparable to the standard quantitative PCR method. Additional target genes associated with antibiotic resistance (*TEM-1, VanA, KPC* and *SHV*) have also been successfully detected *in vitro* and are ready for clinical evaluation. Future development includes multiplexing the sensor to simultaneously detect up to three MDROs target genes, including β-lactamases that hydrolyze β-lactams, the most commonly used antibiotics in clinical settings. This novel sensor platform will be a rapid, economical, point-of-care device with little requirement of reagent handling or technical training.

## 1. Introduction

The lack of effective antibiotic therapies against resistant pathogens leads to increased mortality, increased hospital length of stay, and excessive direct healthcare costs as high as $20 billion/yr.^2, 3^ Antibiotic resistant bacteria such as *Clostridium difficile* and beta-lactam and carbapenem-resistant *Enterobacteriaceae* can colonize the gastrointestinal (GI) tract of patients, and in some cases, can dominate the gut microbiota, causing dysbiosis. This is especially a concern for patients who are immune-compromised (i.e. cancer and transplant patients). In fact, increased abundance of *Enterobacteriaceae* and *Enterococcus spp*. in the gut, many of which are MDROs, can significantly increase the risk of bacteremia with these specific bacteria in cancer patients.^4^ It was shown in pre-clinical models that 1-2 log fold reduction in bacterial colonization significantly decreases dissemination or mortality.^5^ Thus, active monitoring of GI microbiota populations to assist targeted manipulation of the gut immune effectors to decrease bacterial colonization could become standard of care. As part of a national public health surveillance system that tracks changes in the antimicrobial susceptibility of enteric bacteria, the Center for Disease Control (CDC) recommends specialized monitoring to reduce the occurrence of these organisms and related infections (REF?). Monitoring the MDROs in the intestinal tract is therefore critical for providing early warning of emerging infections, identifying resistance patterns, and informing future clinical decisions.

The traditional diagnostic method of culturing MDROs from stool samples is time-consuming (several days), prone to failure since numerous pathogens cannot be cultured, and involves risk of contamination. With the advent of next-generation (NextGen) sequencing technologies (16S rRNA and metagenomic shotgun sequencing), the presence and relative abundance of MDROs can now be detected.^6^ Unfortunately, NextGen sequencing is currently not feasible for clinical use in many settings because samples need to be batched due to high cost, often hundreds of samples at a time, and downstream bioinformatics analysis can be time-intensive. Current diagnostic options include lab-based laborious and complex testing which is expensive and immobile. To identify high-risk patients prone to bacteremia and contain transmission of MDROs, a quick method for determining the “health” of a patient’s gut microbiota would be an invaluable tool. Thus, there is an urgent need to rapidly identify and quantify targeted MDROs, as well as provide a snapshot of the overall microbiota balance in the gut.

Molecular diagnostic assays promise the identification of MDROs at the genus and species levels from stool samples, allowing for accurate and definitive diagnosis. Many polymerase chain reaction (PCR) based protocols have been described that utilize MDRO identification by amplification of bacterial DNA; however, it is difficult to adapt PCR for rapid testing due to its limitations like interferences in biological samples, length of analysis, and high false positive rates resulting from non-specific amplification.^7^ A PCR-free platform is therefore more practical to achieve rapid detection, appropriate treatment decisions, and disease surveillance.

We report a rapid and simple diagnostic assay to monitor MDROs in the lower intestinal microbiome without the use of sequencing or nucleic acid amplification methods. During this study, we demonstrated highly sensitive detection of MDRO gut microbiota targets, increased throughput by simultaneously testing each sample against a panel of target genes and pre-clinical research that demonstrates the capability of our assay in detecting gut microbiome disruption. Our enzymatic assay has detection levels down to attomolar sensitivity with high assay reproducibility. Successful development of the GEMSens (<u>G</u>iner <u>E</u>lectrochemical <u>M</u>icrobial <u>Sens</u>or) platform will lead to many benefits, including early diagnosis of gut microbiome disruption, an infection control-based intervention, and capability to assist advanced microbiome therapeutic methods.

## 2. Materials and Methods

### 2.1 Materials and Equipment

All oligonucleotide (custom capture probes [Catalog #211205107], *KPC* labeling probes as P1, P2, P3, and target DNA of MDRO, *KPC* Long [Catalog #214932680]), *TEM-1, vanA*, and *SHV* sequences were synthesized and purchased from Integrated DNA technologies (IDT), USA, and their sequence information is mentioned in the supplementary material **(Table 4)**. Streptavidin-coated 96 well electrode plates (DRP-96X110STR), screen-printed carbon electrodes (DRP-CONNECTOR96X) and Sync Box (DRP-SYNCONN96X) were purchased from Metrohm USA Inc., Riverview, FL. Blocker BSA (1%) in PBS (Catalog # 37525) were purchased from Thermo Scientific. 3,3’,5,5’-tetramethylbenzidine (TMB) substrate solution was bought from Thermo Fisher (34028). Invitrogen’s 10X PBS Buffer with pH 7.4 (AM9629), along with a 20X PBS-T buffer (228352) from Thermo Fisher were used in all studies. The High Sensitivity Streptavidin HRP was purchased from Thermo Fisher (21130). Poly-HRP conjugate was purchased from Fitzgerald Industries International, Acton, MA. Biotin (Catalog # B4501), H_2_SO_4_ and MgCl_2_, and all other reagents were purchased from Surmodics, Eden Prairie, MN. Invitrogen’s Ultrapure DNase-free, RNase-free DEPC treated water (catalog # 4387937) was used in all studies. The Li-ion powered multichannel potentiostat used for chronoamperometry (Model number: µSTAT4000P, Potential range ±3 V or ±4 V, and Current ranges 1 nA to 10 mA or 100 mA) was obtained from PalmSens BV, Houten, Netherlands.

### 2.2 Methods

#### 2.2.1. Experimental stock preparation

Poly-HRP stock is diluted to 30X in the buffer (1%BSA+1X PBS). Enough stock should be prepared for 100 µL per well (3 or 4 wells). 100 µL of Capture Probe solution is prepared by adding 25 nM of Capture Probe solution into 20mM of MgCl_2_ + 1X PBS buffer, resulting in a final concentration of 25 nM. DNA oligonucleotides were resuspended in PBS buffer and stored at - 20°C as DNA stocks. 100µL of DNA amplification probe mix was prepared for each probe (P1, P2, P3) at a concentration of 25 nM. DNA probe immobilization and target solution were prepared with 20 mM of 1X PBS buffer, including MgCl_2_ to give 100 µL per well. In addition, 1X Phosphate Buffered Saline-Tween (PBS-T) buffer and 1X PBS buffer were prepared as washing solutions from the buffer stock solutions. The poly-HRP80-Streptavidin conjugate was diluted in 1X PBS buffer, including 1% BSA at required dilution rates. In addition, 40 µM of freshly prepared biotin solution was prepared in nuclease free DEPC treated water, to achieve enough solutions for 100 µL per well (3 or 4 wells). H_2_SO_4_ stock solution was diluted with DI water to obtain 0.5 M H_2_SO_4_ solution. DEPC-treated water was used for all other solutions preparation throughout the experiments.

#### 2.2.2. Assay development

Herein, we developed an enzymatic sensing system to detect MDRO genomic DNA in the gut microbiome utilizing a single-stranded capture probe (Cp) and three labeling oligonucleotide probes (P1, P2, and P3), as shown in **Fig. 1**. Briefly, the capture probe was designed to hybridize with a nucleic acid region on single-stranded target DNA. This probe was modified with biotin at its 5′ end to attach to the streptavidin-coated 96-well plate surface. Before adding the capture probe to the wells, pre-wash with 300 μL 1X PBS-T. After the 25 nM of capture probe was immobilized on the plate surface and shaken for 30 minutes at 500 rpm, the sample solution containing single-stranded target DNA molecules was added to the system and was also shaken for 30 minutes at 500 rpm. This study designed synthetic target DNA molecules as approximately 200 bps long for *TEM-1, vanA, KPC* and *SHV*. After hybridization, the wells were washed three times with 1X PBS-T buffer, and each well was treated with 40 μM of freshly prepared biotin solution for 10 min at room temperature (RT) to avoid possible nonspecific bindings. Once biotin is added to the surface of the wells shake for 15 minutes at 500 rpm. 100 μL of the labeling probe mixture, which was previously prepared in one tube to be 25 nM of each P1, P2, and P3 probe, was added to the wells and shaken for 30 minutes at 500 rpm to hybridize along the length of the target DNA. After washing the wells with buffer solution, 30X Streptavidin poly-HRP80 conjugate (prepared with 10X of 1% Blocker BSA solution with 1% PBS) was introduced to the wells and shaken for 30 minutes at 500 rpm to attach to the labeling probes by biotin-streptavidin interaction. After the washing step, 100 μL of room temperature stock TMB was added to the wells. Poly-HRP catalyzes the oxidation of TMB in the presence of hydrogen peroxide. After ∼10 mins, 50 μL of 0.5 M H_2_SO_4_ was added to the system to stop the reaction. Each well was analyzed by UV/Vis spectroscopy (at 450 nm) and chronoamperometry. The oxidized form of TMB can be measured by chronoamperometry once converted to the diimine form in acidic conditions.^8^

**Fig. 1.**
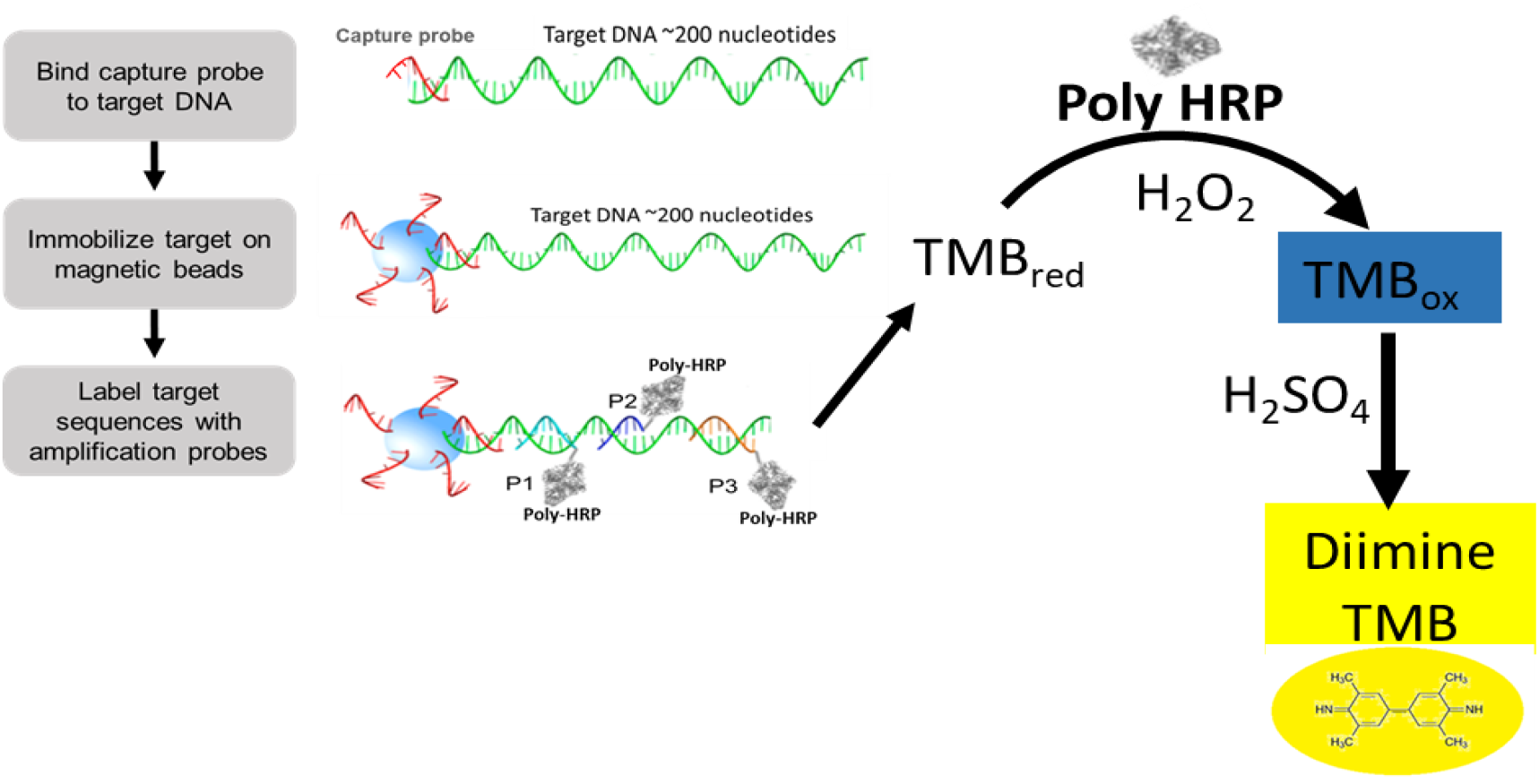
Schematic of DNA detection via an enzyme linked “sandwich” assay. Signal is generated by the target sequence linking poly-horseradish peroxidase (poly HRP) to the magnetic GEMs, which generates an electroactive product from tetramethylbenzidine (TMB) substrate in solution. Figure adapted from Song et. al.^1^

#### 2.2.3. Sensitivity measurements

Spectrophotometric and electrochemical measurements were performed using various concentrations of target DNAs designed separately for the *KPC, SHV, TEM-1* and *vanA* gene regions. Ranging concentrations of target DNA (10 pM down to 10 nM) were added to the wells to hybridize with the 25 nM of accordingly designed capture probes. Control experiments were performed in the absence of the target gene while keeping every other parameter unchanged. For all electrochemical measurements in this research, 70 μL of TMB solution was added onto the electrode surface, followed by amperometric measurements. An operating potential of 125 mV was applied to decay the transient currents to a steady-state value. The time required for this was determined as 30 s in this study. Sharply increasing current implies that TMB reduction has been achieved.

#### 2.2.4. Specificity measurements

The assay’s specificity was evaluated for each target sequence. A complex cocktail mixture of non-target genes was utilized to show that the assay can detect only the targeted gene sequence in vitro PBS buffer as well as in exogenously enriched UTSW’s provided mouse and human stool samples.

#### 2.2.5. Detection in mouse stool samples

Our collaborator, UTSW processed all mouse stool samples. Mouse stool samples were utilized to demonstrate the preclinical utility of GEMSens. Prior to testing mouse fecal samples were exogenously enhanced with E. coli. The fecal specimens of the mice were spiked with 0.01 ng/µL of the targeted genes, *KPC, TEM-1, vanA*, and *SHV*. The genes containing drug resistant elements were tested using synthetic DNA samples and fecal slurry samples. Both exogenously enriched (spiked) and un-spiked samples were tested, followed by the remaining enzymatic steps as described above. These 10 spiked solutions were then processed as typical fecal samples using a phenol-chloroform extraction protocol^9^ to extract all DNA (E. coli and mouse microbiome).

## 3. Results and Discussion

The GEMSens biosensor we designed utilizes a biotinylated probe that binds to the streptavidin modified electrodes to capture select target DNA sequences. Once captured, sequence-specific label probes (P1, P2, and P3) attach to the target DNA sequence (**Tables 1-4, Fig. S1-S2**). Poly-HRP80, an enzyme attached to the labelling probes, then catalyzes the oxidation of 3,3′,5,5′ - Tetramethylbenzidine (TMB) in the presence of hydrogen peroxide (H_2_O_2_) **(Fig. 1)**. The oxidized form of TMB is subsequently reduced by chronoamperometry once it is converted to its diimine form in acidic conditions **(Fig. 2)**. Because the TMB molecule is oxidized by poly-HRP80, the chronoamperometric analysis focuses on the cathodic current that is produced when the TMB is reduced. The peaks observed during the reduction step are proportional to the amount of TMB in the solution, and thus the targeted DNA. **Figures S3-S5** show the capture probe saturation, probe conc. optimization for sensitivity improvement with gel electrophoresis and assay control study.

**Fig. 2.**
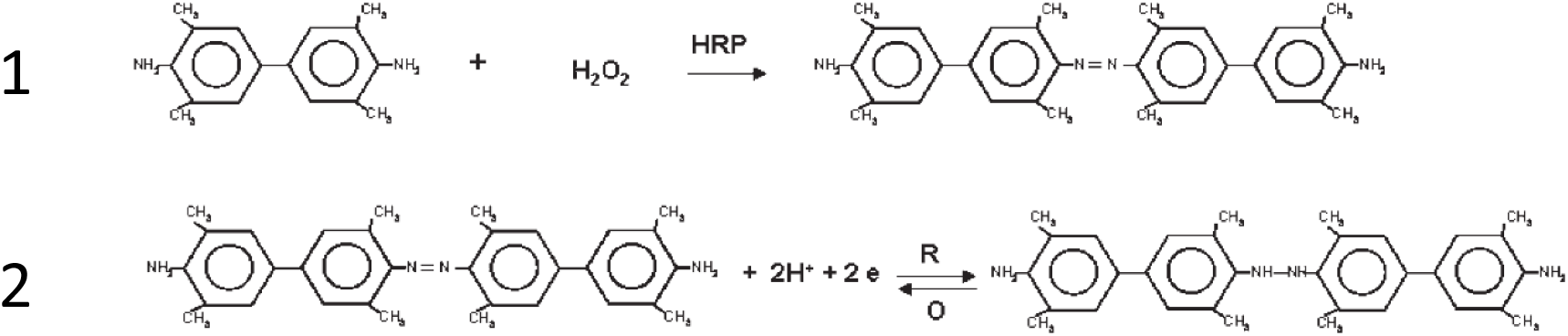
Reaction equation for the oxidation of TMB by HRP and the subsequent reduction of TMB by electrochemical analysis. Adapted from Liu et al.^5^

### 3.1 Enzyme Linked Assay for Electrochemical Detection

As shown in **Fig. 3**, we observed a significant amount of signal enhancement in presence of 0.1 fM, 10 fM, 100 fM and 1000 fM concentration setting compared to control (absence of target) experiment. We validated this signal enhancement in 96-well plates both colorimetrically through UV-Vis at 450 nm **(Fig. 3A)** and electrochemically using chronoamperometric analysis **(Fig. 3B-C)**. In **Fig. 3A** inset, in absence of target, the “sandwich” assay showed a very light blue color (background) that transitioned to light to dark yellow color in the presence of H_2_SO_4_. When the target was present, a bright and dark blue color was observed due to poly-HRP triggered oxidation of TMB, which transitioned to bright orange when acid was added. In order to normalize the intra-assay or plate-to-plate variances, we normalized the data with background subtraction. The standard curve with R^2^=0.9933 value from electrochemical amperometric peak analysis shows excellent linearity and correlation between peak current and concentration of target DNA proportional to oxidize TMB through HRP interaction to provide high sensitivity. To track and assess the sensitivity, it is imperative to evaluate the sensitivity of the electrochemical technique. The standard three sigma rule can consider the limit of detection (LOD) to be calculated using the formula LOD = 3.3 σ/s. ^10-12^ Where σ is the standard deviation, and *s* is the slope obtained through the calibration plot. The LOD of *TEM-1* DNA was estimated to be 0.1 fM.

**Fig. 3.**
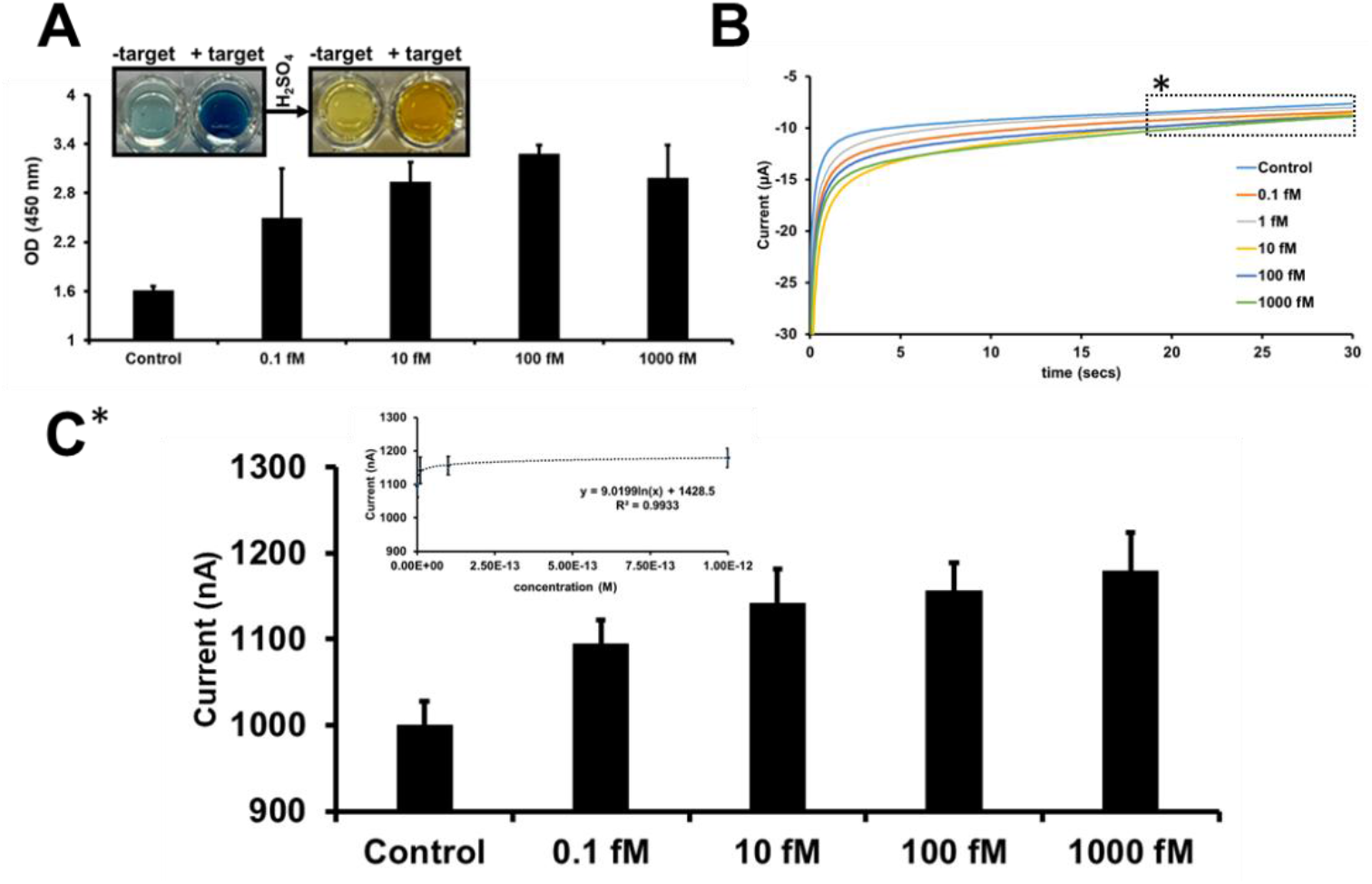
Enzyme linked assay for ultra-sensitive *TEM-1* DNA detection. A. Spectroscopic sensitivity measurement (color transition due to TMB-HRP interaction related to TEM-1 conc.) at 450 nm. B. Amperometric sensitivity measurements of TEM-1 target. C. A graphical concentration dependent calibration curve of amperometric signal (R^2^=0.9933, inset figure) (^*^averages between 20 secs to 30 secs data points) at 125 mV. The last 10 secs data points were considered as stable signal (plateau phase) readout for individual experimental concentration. All experiments were performed in triplicate.

As shown in **Fig. 4**, we observed a significant amount of signal enhancement in presence of 0.01 fM, 0.1 fM, 10 fM and 100 fM concentration setting compared to control (non-target) experiment. We validated this signal enhancement in 96-well plates both spectroscopically at 450 nm (data not shown) and electrochemically using chronoamperometric analysis (**Fig. 4**). The experimental data shows single data set but originally performed in triplicate (n=3). This signal enhancement was observed due to poly-HRP triggered oxidation of TMB, which transitioned to bright orange when acid was added.

**Fig. 4.**
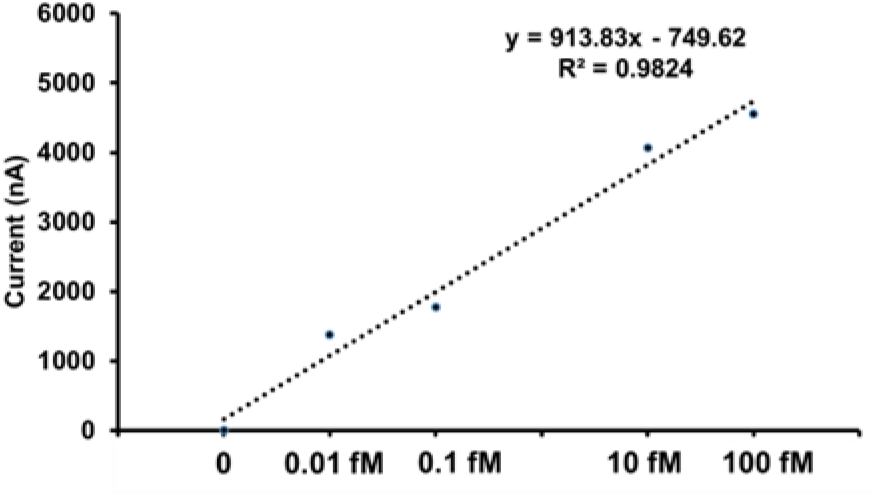
Sensitivity test. Improvement of enzyme linked assay for ultra-sensitive TEM-1 DNA detection. Amperometric sensitivity measurements of *TEM-1* target. A graphical concentration dependent calibration curve of amperometric signal (R^2^=0.9824) (^*^averages between 20 secs to 30 secs data points, data not shown) at 125 mV. The last 10 secs data points were considered as stable signal (plateau phase) readout for individual experimental concentration.

In absence of target, the “sandwich” assay showed a very light blue color (background) that transitioned to light to dark yellow color in the presence of H_2_SO_4_. When the target was present, a bright and dark blue color was observed. To normalize the intra-assay or plate-to-plate variances, we normalized the data with background subtraction. The standard curve with R^2^=0.9824 value from amperometric peak analysis shows excellent linearity and correlation between peak current and concentration of DNA proportional to oxidized TMB through HRP interaction to provide high sensitivity (LOD= 10 aM) using the formula LOD = 3.3 σ/s. ^10-12^

Next, we evaluated the specificity of the enzyme-linked assay for sensitive detection of the *TEM-1* target within complex mixtures containing the non-target genes *SHV* and *vanA*. Both UV–Vis (data not shown) and electrochemical amperometric measurements (**Fig. 5**) confirmed that the assay selectively detected the intended target, even in the presence of competing sequences. This was demonstrated using two cocktail conditions: cocktail 1 (*TEM-1: vanA: SHV* = 1:0.01:0.001) and cocktail 2 (1:1:1).

**Fig. 5.**
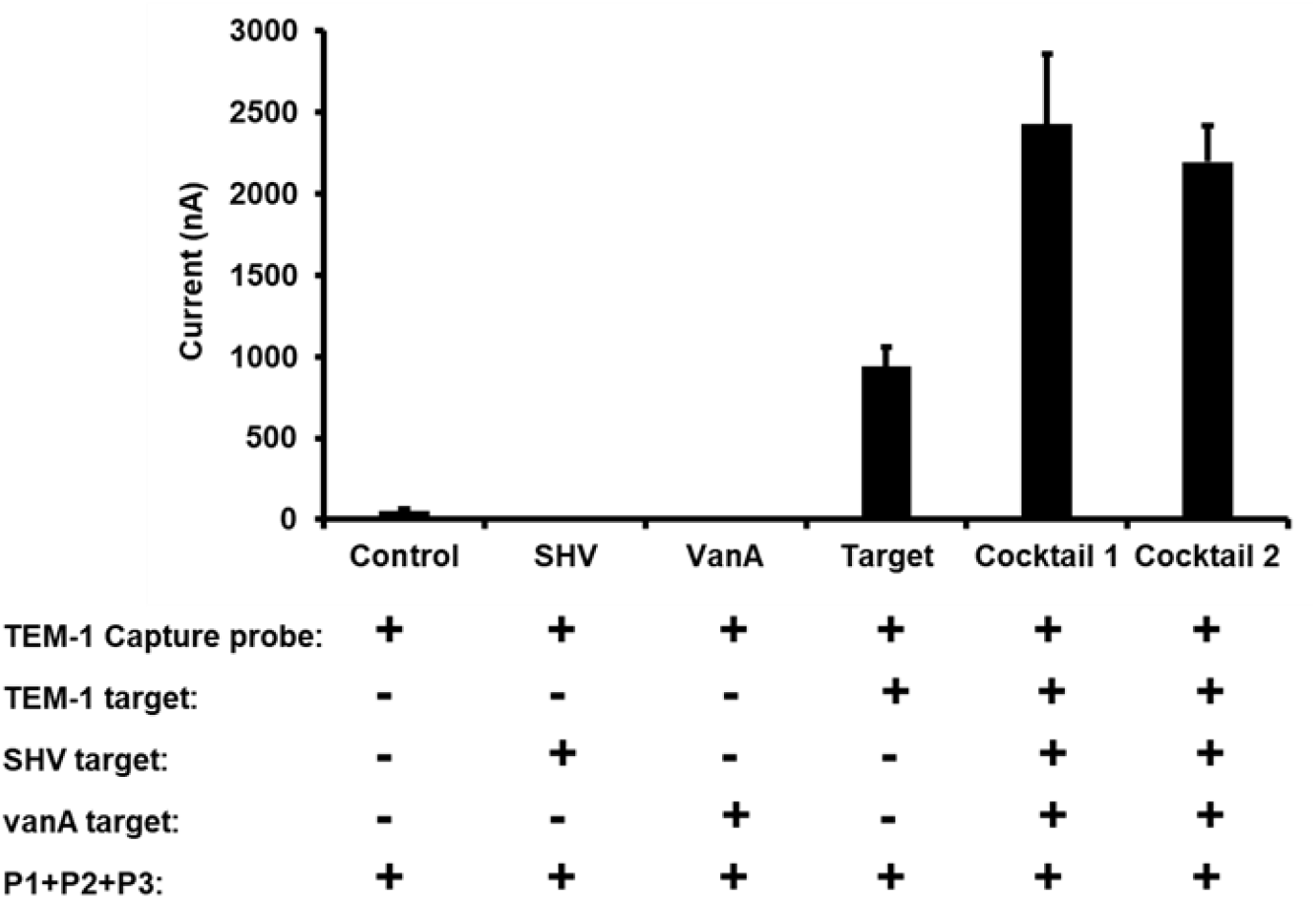
Specificity test. (control without *TEM-1* target, non-targets such as 10 nM of *SHV* ssDNA and a cocktail-1 mixture of 3 targets; 5 nM of *TEM-1*, 0.05 nM of SHV and 0.005 nM of vanA ssDNA target, cocktail-2 with the ratio of each 1:1:1). Biotin-labelled sequence specific amplification probes (P1, P2 and P3) were present in all settings. Electrochemical specificity confirms signal enhancement from both cocktail 1 and 2 mixture due to presence *TEM-1* target than other group where TEM-1 target is absent.

To improve reproducibility, we implemented a standardized sequential workflow, including the use of the same batch of plates and electrodes, freshly prepared buffers, and fixed incubation times. These measures minimize standard deviation and reduce assay-to-assay variability in signal readout.

This set of data shown in **Fig. 4 and 5** indicates that this new enzyme-linked alternative assay has high potential for ultra-sensitive (attomolar level) and rapid (less than 30 minutes) detection of target genes of interest in complex biological matrix with high specificity. The use of 96-well plates provides high throughput and ease of integration with other components of our electrochemical assay (**Figure S6**). We have repeated this set of experiments multiple times to verify the reproducibility and finally able to successfully incorporate an inter-assay variation cofactor with background correction (or normalization) to minimize the plate-to-plate signal variation.

### 3.2 Detection of MDRO Genes (KPC, SHV and vanA) in Fecal Samples

We evaluated the enzymatic assay using fecal DNA extracts described in Section 2.2.5. Electrochemical signal enhancement was observed for *KPC, SHV*, and *vanA* gene targets in mouse fecal samples (total DNA input ∼0.01 ng/µL), as shown in **Fig. 6**. Detectable signals were obtained even for the lowest input condition (A1; fecal slurry spiked with 10^1^ cells), where responses for *KPC* and *vanA* exceeded the control blank (commensal bacteria only), indicating high analytical sensitivity of GEMSens.

**Fig. 6.**
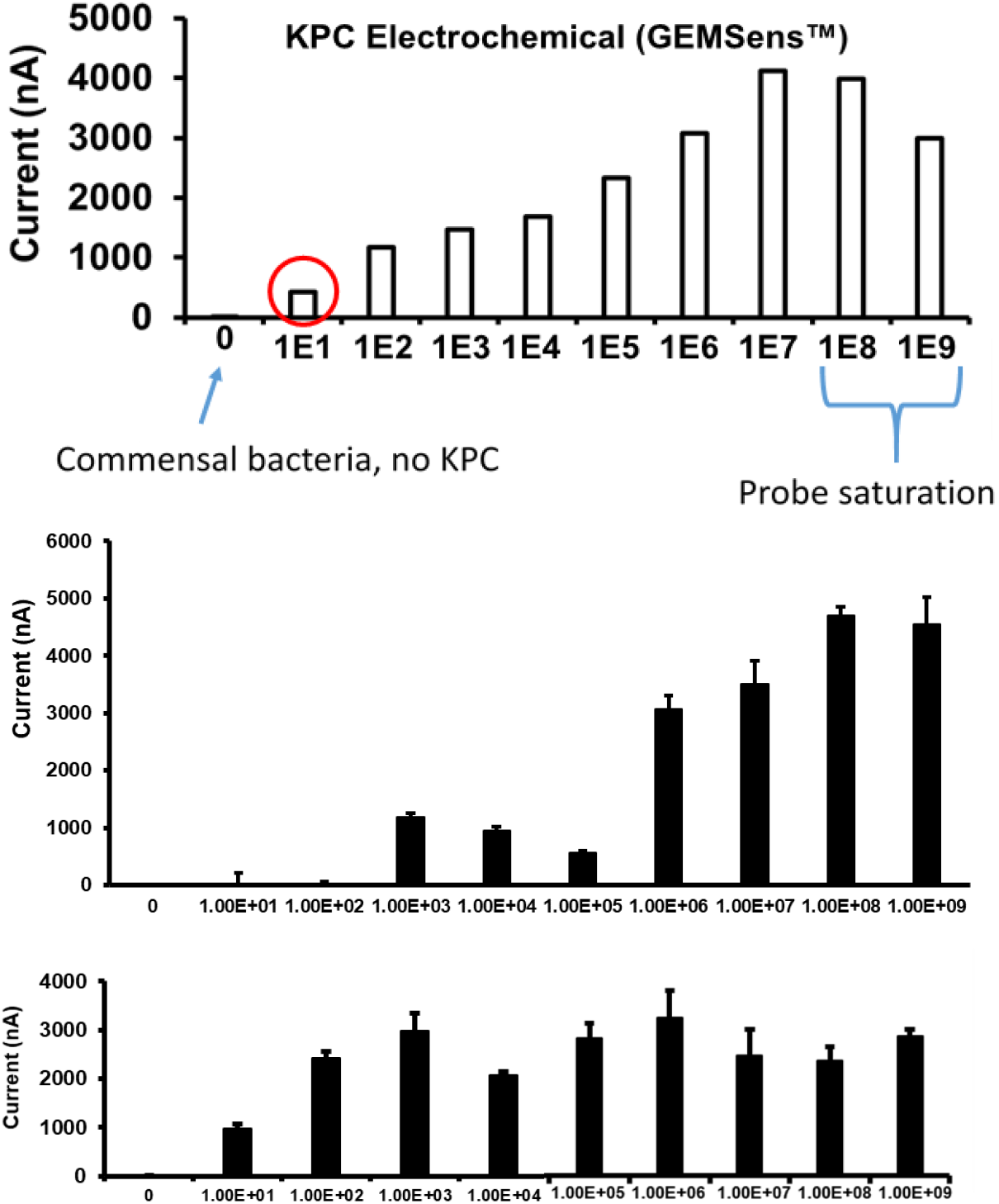
**A**. KPC; **B**. *SHV* and **C**. vanA gene detection in mouse fecal extracts with Giner’s enzymatic electrochemical assay. All experiments were performed in triplicate except KPC.

For *KPC* and *SHV* (**Fig. 6A–B**), signal intensities increased with target concentration at lower inputs but approached a plateau at higher levels (≥10^7^ cells), consistent with saturation of probe binding on magnetic beads. Across A1–A9, the signal progression was not strictly linear (particularly for *SHV* and *vanA*), although all target-containing samples produced signals above background. Background correction was applied to mitigate well-to-well variability.

A similar trend was observed for *vanA* (**Fig. 6C**), with an approximately linear response at lower concentrations (A1–A3), followed by reduced proportionality at higher inputs. No measurable signal was detected in PBS or un-spiked fecal controls.

Overall, these results demonstrate that the assay enables reliable detection of target genes down to ∼10 CFU/g (A1, highlighted in **Fig. 6**) for KPC and *vanA*. While the response is not fully quantitative across the entire range—likely due to probe saturation and electrode-to-electrode variability-the assay provides sensitive and reproducible qualitative detection in complex fecal matrices (**Figs. S7–S9**).

### 3.3 Human Sample Testing

After optimization of the assay, we evaluated its performance using archived human stool samples obtained from UTSW. Genomic DNA extracts were spiked with varying amounts of four target genes. As an example, **Fig. S7** shows results from a sample spiked with a high concentration of *E. coli* (strain AR 0546) harboring the KPC gene (∼10^9^ CFU/g stool). As expected, a strong signal was observed with the *KPC*-specific probe (**Fig. 7**), confirming successful target detection in a complex clinical matrix.

**Fig. 7.**
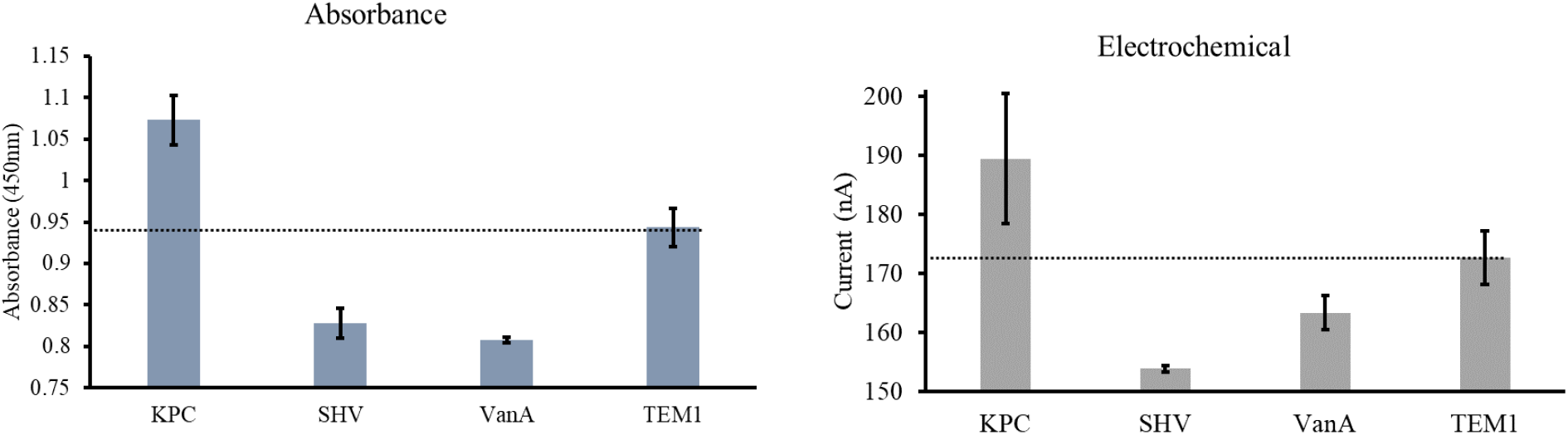
Clinical human sample testing with GEMSens-E™ (Giner) assay. The sample had a high concentration of the KPC gene added to the sample by spiking stool samples with 10^9^ CFU/g of bacteria. This was verified with Giner’s GEMSens-E^™^ results.

Low-level signals observed for non-target probes (*SHV, vanA*, and *TEM-1*) are unlikely to arise from direct probe–target complementarity, as probes were designed against distinct regions and show conservation region primarily within, but not across, gene families (**Figs. S1–S2**). Instead, these background responses are more likely attributable to nonspecific interactions and matrix effects associated with stool-derived samples. Importantly, target-specific signals remained clearly distinguishable after background correction.

## 4. Conclusion and outlook

During this study, we developed the MDRO detection technology and the proof of feasibility of the assay.^13^ In collaboration with the UTSW team, we successfully identified DNA probe sequences to target four drug resistant bacteria genes. We successfully developed novel enzyme-based electrochemical assay (GEMSens) that shows attomolar sensitivity (LOD= 10-100 aM), and the sample to result in turnaround time is less than an hour. The enzymatic assay was the primary focus of the program due to significantly increased speed and sensitivity.

Prior to human sample testing, we evaluated the pre-clinical utility of GEMSens through testing at least 4 genomic culture samples and ∼90 mouse fecal samples spiked with target MDRO genes (*TEM-1, vanA, KPC* and *SHV*). GEMSens sensor was capable of MDRO genomic DNA detection down to 10 colony-forming units (CFU)/g in mouse fecal samples. Overall, the GEMSens assay is reproducible, highly sensitive, specific (no false positive/negative signal confirmed with a cocktail of non-target genes) and validated (cross evaluated with *the in-house* optical assay). Additionally, using the high-throughput assay, we can process 8 distinct samples per 96-well plate within 45 mins time frame. The adaptation of the GEMSens assay for 96-well plate testing provided a strong foundation for a rapid, cost-effective, and high-throughput test to monitor MDRO domination in the gut microbiome. Finally, our initial human clinical tests have shown promise for GEMSens to be a useful pre-clinical to clinical tool in the rapid detection of drug-resistant bacteria in healthcare facilities.

## Data Availability

All data produced in the present work are contained in the manuscript

## Author Contributions

A.A.A and M.R.^a^ conceived the study and designed the experiments. M.R.^a^ performed the bioinformatics, identified, down selected, and designed all the oligos in collaboration with E.T. M.R.^bc^ and A.K. M.R.^a^ performed the absence test, sensitivity, specificity, and detection experiments in mouse fecal extracts and human stool samples. M.R.^a^, M.S. and A.A.A wrote the manuscript.

## Acknowledgements

M.R.^a^, M.S., M.R.^bc^, E.T. A.K. and A.A.A acknowledge the funding support from Department of Health and Human Services, Centers for Disease Control and Prevention (CDC) through an SBIR Phase II contract (Contract # 200-2017-95643). We would like to thank Andrew Weber, Saylor Bower, Eric Kilbourn, Mourad Manoukian, and Claire Tefler for intellectual feedback, helpful discussions, and guidance.

