## Supplementary Information for "An Ultrasensitive and Novel Gene Sensor to Monitor Gut Microbiome"

**Probe Design for Antibiotic Resistant Genes**

**Table 1.** *Probe design for Klebsiella pneumoniae carbapenemase (KPC).* KPC are classified as Group A carbapenemases which confer reduced susceptibility to carbapenem antibiotics. These genes are usually found on plasmids and therefore have greater potential for dissemination.

| Gene | | KPC | |
|---|---|---|---|
| QPCR FOR | ATGCGCGCGATACCTCATC | QPCR REV | GGTCGTGTTTCCCTTTAGCC |
| Sequence 1 | ATGCGCGCGATACCTCATCGCCGCGCGCCGTGACGGAAAGCTTACAAAAACTGACACTGGGCTCTGCACTGGCTGCGCCGCAGCGGCAG CAGTTTGTTGATTGGCTAAAGGGAAACACGACC | | |
| PROBE 1 | GACGGAAAGCTTACAAAAACTGACACTGGGCTCTGCACTGGCTGC | | |
| QPCR FOR | TTACGGCAAAAATGCGCTGG | QPCR REV | CGGCGGCGTTATCACTGTAT |
| Sequence 2 | TTACGGCAAAAATGCGCTGGTTCCGTGGTCACCCATCTCGGAAAAATATCTGACAACAGGCATGACGGTGGCGGAGCTGTCCGCGGCCG CCGTGCAATACAGTGATAACGCCGCCG | | |
| PROBE 1 | GGAAAAATATCTGACAACAGGCATGACGGTGGCGGAGCTG | | |

**Table 2.** *Probe design for ESBL SHV.* SHV is one of the extended spectrum beta lactamases (ESBL) that is commonly found in ampicillin resistant E. coli and K. pneumoniae strains in the clinic.

| Gene | | SHV | |
|---|---|---|---|
| QPCR FOR | ATGCGTTATATTCGCCTGTG | QPCR REV | TTTCTATCATGCCTACGCGG |
| Sequence | ATGCGTTATATTCGCCTGTGTATTATCTCCCTGTTAGCCACCCTGCCGCTGGCGGTACACGCCAGCCCGCAGCCGCTTGAGCAAATTAAAC AAAGCGAAAGCCAGCTGTCGGGCCGCGTAGGCATGATAGAAA | | |
| PROBE 1 | TATATTCGCCTGTGTATTATCTCCCTGTTAGCCACCCTGCCGCTG | | |
| PROBE 2 | TTGAGCAAATTAAACTAAGCGAAAGCCAGCTGTCGGGCCGCGTAGGCATG | | |

**Table 3.** *Probe design for vanA.* vanA confers resistance to vancomycin, a drug commonly used to treat gram-positive infections. Vancomycin-resistant enterococci such as E. faecium are classified as a "serious" threat according to the CDC

| Gene | | *vanA* | |
|---|---|---|---|
| QPCR FOR | ACGAGCCGTTATACATTGGAAT | QPCR REV | CGGCGAGAGTACAGCTGAAT |
| Sequence | ACGAGCCGTTATACATTGGAATTACGAAATCTGGTGTATGGAAAATGTGCGAAAAACCTTGCGCGGAATGGGAAAACGACAATTGCTAT TCAGCTGTACTCTCGCCG | | |
| PROBE 1 | AATTACGAAATCTGGTGTATGGAAAATGTGCGAAAAACCT | | |
| PROBE 2 | TATGGAAAATGTGCGAAAAACCTTGCGCGGAATGGGAAAACGACAATTGC | | |

**Table 4.** Oligonucleotide sequences for TEM-1, vanA, SHV, and KPC probe testing.

| Name | Length (bases) | Sequence |
|---|---|---|
| *TEM-1 probe* | 40 | 5'-/5Biosg/GGT TGA GTA CTC ACC AGT CAC AGA AAA GCA TCT TAC GGA T-3' |
| *TEM-1 target* | 40 | 5'-ATC CGT AAG ATG CTT TTC TGT GAC TGG TGA GTA CTC AAC C-3' |
| *vanA probe* | 40 | 5'-/5Biosg/AAT TAC GAA ATC TGG TGT ATG GAA AAT GTG CGA AAA ACC T-3' |
| *vanA target* | 40 | 5'- AGG TTT TTC GCA CAT TTT CCA TAC ACC AGA TTT CGT AAT T -3' |
| *SHV probe* | 45 | 5'-/5Biosg/ TAT ATT CGC CTG TGT ATT ATC TCC CTG TTA GCC ACC CTG CCG CTG - 3' |
| *SHV target* | 45 | 5'- CAG CGG CAG GGT GGC TAA CAG GGA GAT AAT ACA CAG GCG AAT ATA -3' |
| *KPC probe* | 40 | 5'-/5Biosg/ GGA AAA ATA TCT GAC AAC AGG CAT GAC GGT GGC GGA GCT G-3' |
| *KPC target* | 40 | 5'- CAG CTC CGC CAC CGT CAT GCC TGT TGT CAG ATA TTT TTC C -3' |

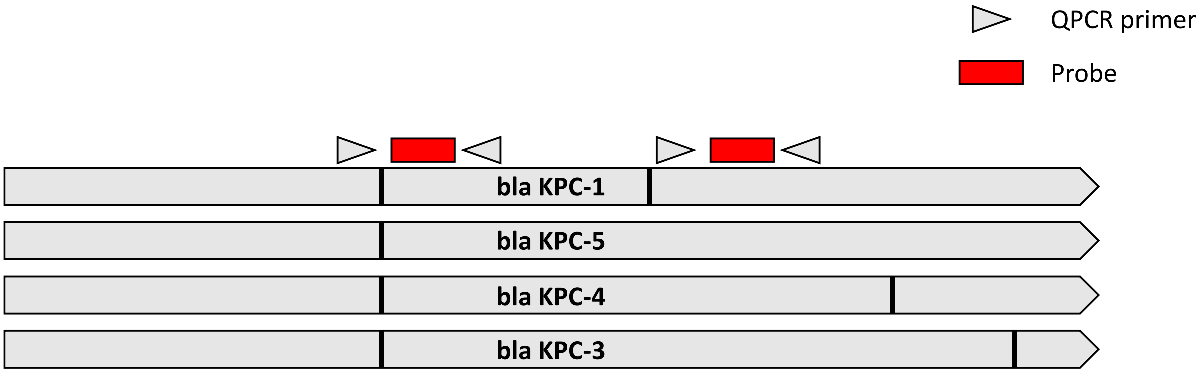


**Fig. S1**. Figure shows the alignment of bla$_{KPC}$ genes (grey arrows). The black lines in the gray arrows are single nucleotide polymorphisms.

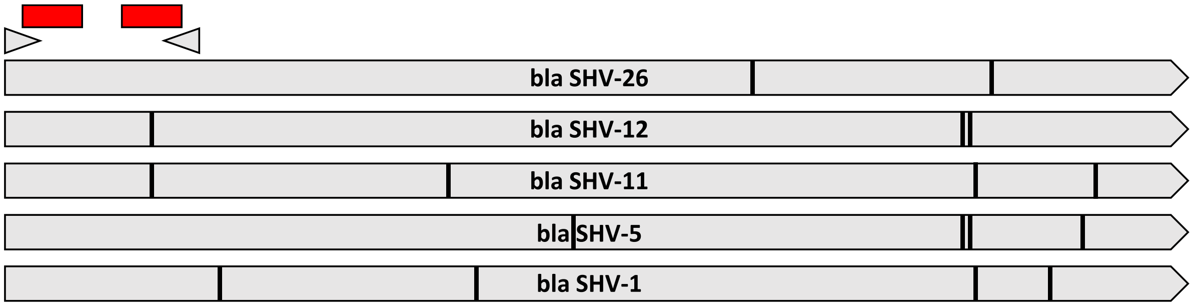


**Fig. S2**. Figure shows the alignment of bla$_{SHV}$ genes for the strains ordered from the CDC AR isolate bank.

### Assay Control Study

Our team optimized the capture probe saturation in **Fig. S4A** shows 5 nM of capture probe (Cp) produces the highest signal to noise ratio. Both 100 nM and 50 nM of capture probe concentrations have high background signals. Probe concentrations below 5 nM also do not yield sufficient signal.

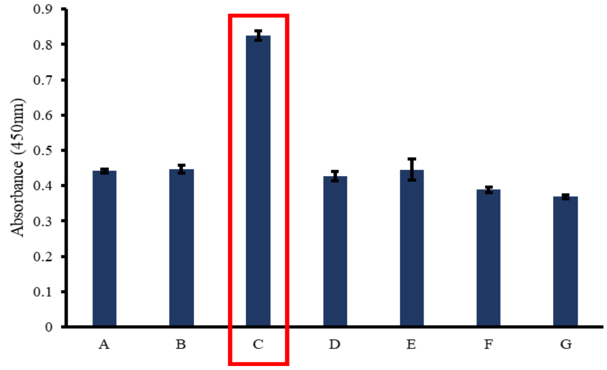


| | Capture Probe | Amplification Probe | Target |
|---|---|---|---|
| **A** | X | | |
| **B** | X | X | |
| **C** | X | X | X |
| **D** | | X | |
| **E** | | X | X |
| **F** | X | | X |
| **G** | | | X |

**Fig. S3.** A control study with multiple combinations of capture and amplification probes to identify any false signal in the assay. As outlined, the only sample that should show a signal is sample C as it is the only sample with all probes and target necessary to produce a signal.

The representing colorimetric images as well as UV-Vis data measured at 450 nm are shown in **Fig. S4A**. The graphical representation of electrochemical measurement is shown in **Fig. S4B**.

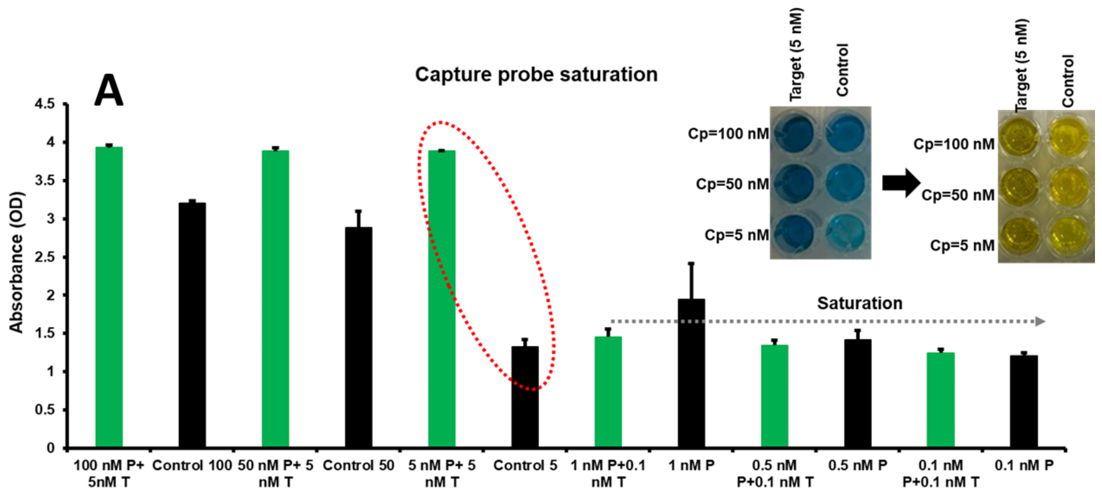


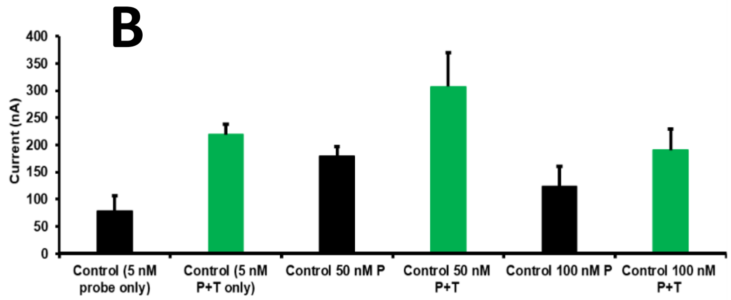


**Fig. S4**. **(A)** Capture probe saturation studies (Cp=100 nM to 0.1 nM) to reduce background signals. Inset. Colorimetric image.

**(B)** Electrochemical signals of capture probe saturation.

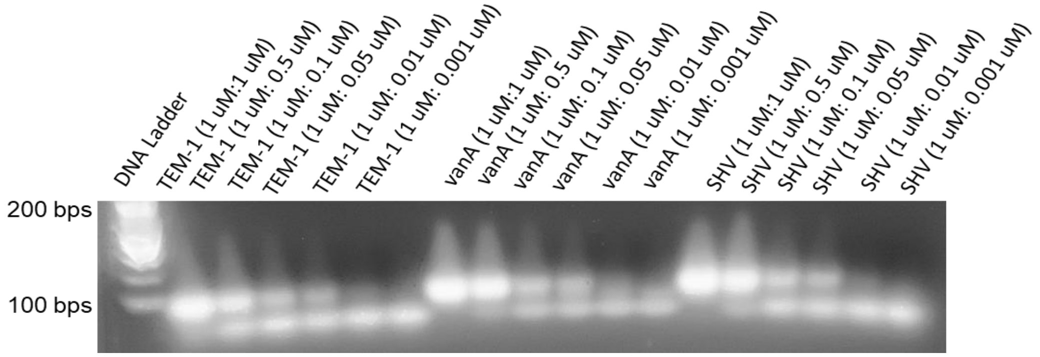


**Fig. S5.** Gel Electrophoresis Data. Optimization of probe concentration for sensitivity improvement.

**Multiplexing the Electrochemical Enzymatic Assay.**

One of the genes of primary focus in this study was *KPC*. We conducted the assay with synthetic KPC gene in concentrations between 10 pM and 10 nM. A sample from end results was scanned with a carbon 96 well plate, and another sample with carbon single electrodes. **Fig. S6** shows an example of the scan shapes that are collected with the 96 well plate and single electrodes. While both have similar shapes, the noise in the 96 well plate scan is noticeably higher than the single electrode scans.

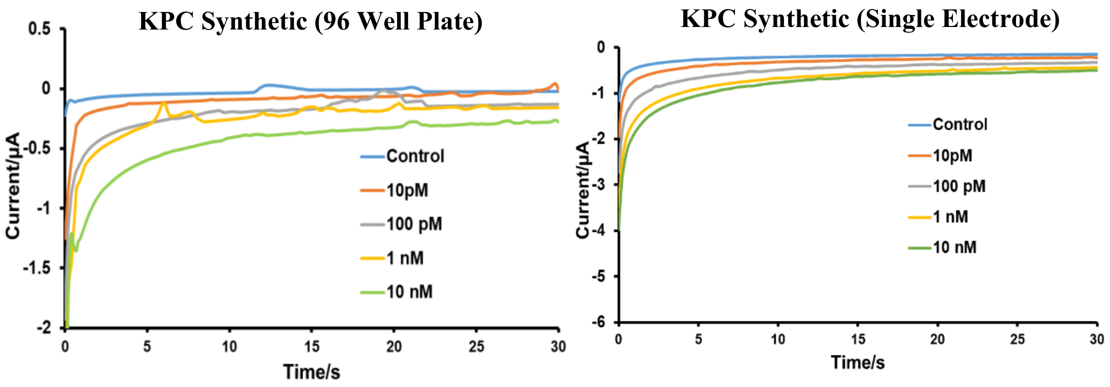


**Fig. S6.** The scans collected using the Metrohm DropView software for a synthetic *KPC* gene target. The 96 well plate scans have significantly more noise than the single electrodes

**Detection of MDRO genes in murine fecal samples.**

Our team has tested the enzymatic assay against the mouse fecal samples provided by UTSW. We observed electrochemical signals in *KPC, TEM-1,* and *SHV* gene spiked fecal samples. The GEMSens assay also tested mouse fecal samples spiked with the *TEM-1* gene. The results can be seen in **Fig. S8** which compares the qPCR results (**Fig. S8A**) to the carbon 96 well plate (**Fig. S8B**)

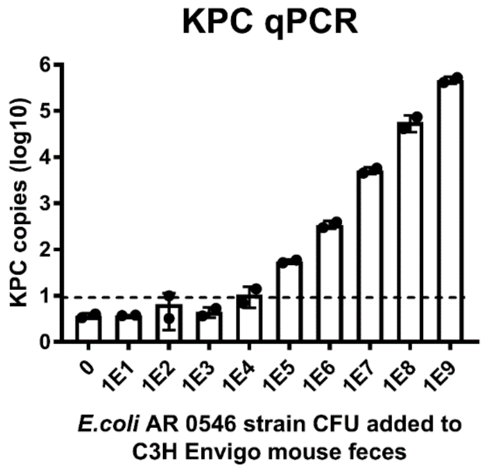


**Fig. S7**. Dilutions of E. coli strain AR 0546 (containing *KPC* gene) were added to mouse feces and total gDNA was purified from each dilution. ‘0’ stands for no E. coli added to the feces. QPCR was performed in order to assess the detection limit above which we can positively identify the KPC gene in the sample. The dotted line represents the detection limit for this assay. The graph represents data from two biological replicates.

and carbon single electrode (**Fig. S8C**) electrochemical results. The *TEM-1* gene shows a general trend of increasing current with increasing gene concentration; however, the assay needs refinement before making a calibration curve for quantitative analysis. There are significant results with gene concentrations as low as 10 CFU/g which is much lower than qPCR's ability to detect.

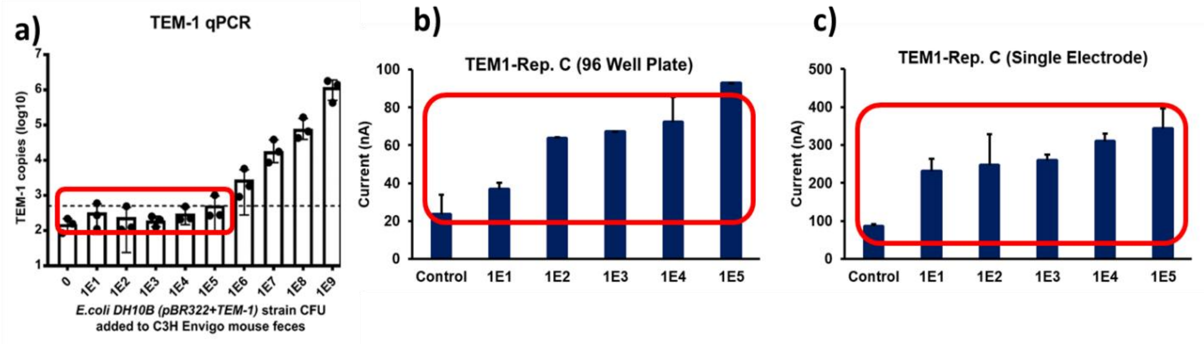


**Fig. S8.** Data collected with qPCR from mouse fecal samples spiked with the *TEM-1* gene **(a)** compared to results from the GEMSens 96 well plate **(b)** and single electrode **(c)**. The results show no signal below $10^6$ CFU for qPCR, but 10 CFU detection for our assay.

**Fig. S9.** shows the full set of data collected with the fecal samples spiked with the *KPC* gene compared to the qPCR results with the same gene (**Fig. S9a**). The samples were scanned with both the carbon electrode 96 well plate (**Fig. S9b**) and single carbon electrodes (**Fig. S9c**). The samples also had spectrophotometry data collected to confirm color change differences (**Fig. S9d**). All three methods of analysis were able to produce significant results with fecal sample gene concentrations as low as 10 CFU/mL and agreed in trend very closely. GEMSens™ assay can detect the presence of MDRO genes in fecal samples substantially lower than qPCR.

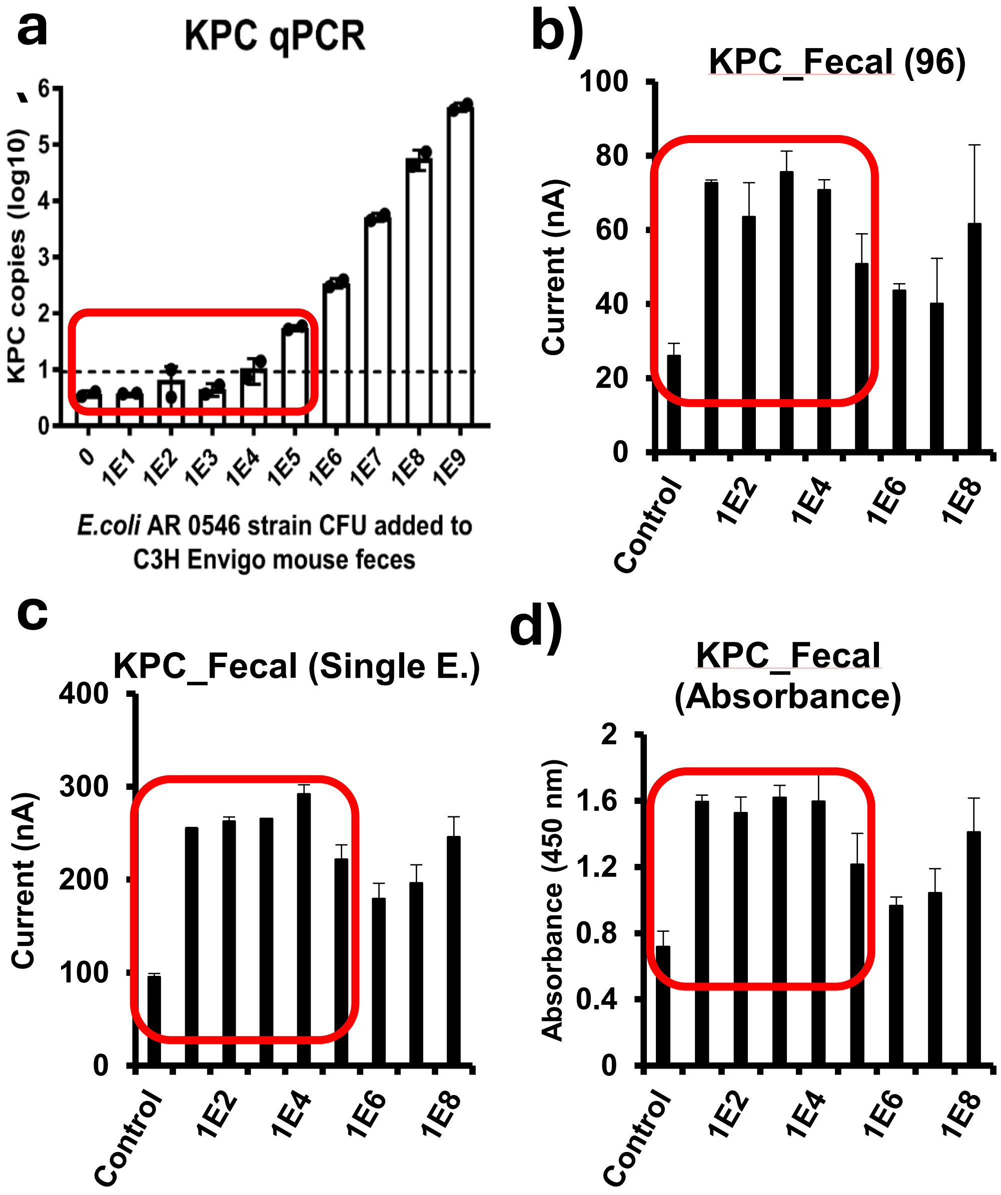


**Fig. S9.** qPCR results **(a)** compared to the GEMSens-E$^{TM}$ results obtained with carbon 96 well plate **(b)**, single carbon electrodes **(c)**, and spectrophotometry **(d)**. The results show significant increase in signal when *KPC* gene is present with a detection limit lower than qPCR.
